# Memory Intraindividual Variability and its Association with Hippocampal Integrity in Older Latino Adults Without Dementia

**DOI:** 10.64898/2026.10.15.26365658

**Authors:** Amanda I. Gonzalez, Catarina Tristão-Pereira, Lusiana Martinez, Nadeshka J. Ramirez, George Aquilino Silva, Jorge Alcina, Jairo E. Martinez, Averi Giudicessi, Vivian Ku, Meredith C. Rowe, Liliana A. Ramirez-Gomez, Daniel G. Saldana, Yakeel T. Quiroz

## Abstract

**Introduction:** Cognitive intraindividual variability (IIV), a measure of fluctuation in performance across time or tasks, is an emerging marker of cognitive decline. However, little is known about within-domain IIV in Latino adults. To address this gap, we examined memory IIV and hippocampal integrity in older Latino individuals.

**Methods:** Sixty-seven participants from the Boston Latino Aging Study underwent cognitive testing and MRI. Linear regressions assessed whether memory IIV was associated with hippocampal volume and hippocampus-cingulum tract fractional anisotropy. Additional analyses evaluated associations using mean memory performance as the outcome.

**Results:** Higher mean performance was associated with greater hippocampal volume (*β*=0.24, *p*=.03), while memory IIV was not (*β*=0.028, *p*=.84). Higher memory IIV was associated with lower fractional anisotropy in the hippocampus-cingulum tract (*β*=-0.34, *p*<.01), while mean performance was not (*β*=-0.07, *p*=.52).

**Discussion:** Memory IIV may be more closely linked to hippocampal white matter, while mean memory performance may better reflect hippocampal volume.

## 1. Introduction

Cognitive intraindividual variability (IIV) is a psychometric measure of fluctuations in an individual’s performance over time or across tasks. Higher IIV, or dispersion (e.g., IIV across cognitive domains), reflects greater inconsistency in performance. Research on IIV is based on theories linking variability in performance to reduced neurological functioning, potentially indicating risk for future decline prior to compromised neuropsychological total scores^1–3^. Recent research has explored IIV as a possible marker of early cognitive decline and neurodegeneration. Specifically, higher IIV has been shown to capture early white matter alterations and proteinopathy characteristic of Alzheimer’s disease (AD) pathology, such as accumulation of amyloid-*β* and tau^4,5^. There is also evidence to suggest that higher IIV indicates higher risk for conversion from normal cognition to mild cognitive impairment (MCI) and dementia, as well as for increased functional burden^5–8^. Earlier detection of AD and related dementia (ADRD) risk may allow for timely intervention and access to preventative strategies, potentially altering disease courses.

Memory change is particularly relevant for the early detection of ADRD. Memory decline is among the earliest symptoms of individuals with dementia, highlighting the importance of studying early memory changes for the early detection of ADRD^9^. The hippocampus is the most well-established brain structure implicated in memory, responsible for memory consolidation and retrieval^10^. Hippocampal function often becomes compromised by disease pathology in AD, making neuroimaging markers of hippocampal integrity critical in monitoring disease risk and progression^11^. The cingulum bundle is a prominent white matter pathway that projects to the hippocampus and is involved in episodic memory^12^. Previous research provides evidence that the hippocampus-cingulum white matter tract is also compromised in AD and associated with greater tau burden^13,14^. Nevertheless, few studies have explored neural correlates of IIV specifically within the domain of memory. One such study found that those with MCI had higher IIV across memory tasks compared to controls and that it was a unique predictor of cognitive status beyond performance accuracy^15^. Similarly, a study examining longitudinal relationships found that higher memory IIV was associated with greater amyloid-*β* plaques, greater neurofibrillary tau tangles, and worse cognitive status at follow-up^5^. While limited, these findings indicate that IIV within the domain of memory may be a sensitive indicator of cognitive decline.

Despite facing higher rates of ADRD, Latino adults continue to be underrepresented in cognitive aging research^16^. Specifically, early memory changes and neuroimaging correlates remain poorly understood in Latino adults. One previous study exploring IIV among Hispanic adults characterized associations between sociodemographic factors and cognitive dispersion across tasks on the Spanish-language NIH Toolbox Cognition Battery. Findings showed that dispersion, calculated as the intraindividual standard deviation (ISD) for crystallized tests (i.e., vocabulary and word-reading tests) was significantly associated with sex, income, and whether participants were educated domestically^17^. However, little is known regarding associations among IIV and neuroimaging markers in Spanish-speaking Latino individuals. Given the lack of literature on memory IIV within this population, we sought to characterize associations between memory IIV and demographic variables such as age, sex, education, and cognitive status (Aim 1). We also aimed to examine the associations between memory IIV and hippocampal integrity, measured as hippocampal volume and fractional anisotropy in the hippocampus-cingulum tract (Aim 2). Lastly, we sought to examine the associations between mean memory performance and measures of hippocampal integrity, to assess whether memory IIV may offer any additional information beyond mean performance (Aim 3).

## 2. Methods

### 2.1 Participants

We collected data from the Boston Latino Aging Study (BLAST), an ongoing community-based longitudinal study of aging launched in 2021. The sample included 67 participants with complete MRI and neuropsychological data available. Study inclusion criteria included being at least 55 years of age, self-identifying as Latino/a, residing in the Greater Boston Area, and being either monolingual in Spanish or bilingual in Spanish/English. Spanish-speaking participants were included regardless of cognitive status, with cognitive impairment classified as a Mini-Mental State Examination (MMSE) total score of less than 24^18^. Study exclusion criteria included any existing dementia diagnosis, history of chronic major psychiatric disorders or psychosis, as well as medication changes within 30 days prior to study completion. Participants provided sociodemographic information, underwent neuropsychological testing in Spanish, and a brain magnetic resonance imaging (MRI) scan. All participants provided informed consent, and study procedures were approved by the Mass General Brigham Human Research Committee (MGHRC), the institutional review board overseeing Mass General Brigham and affiliated institutions.

### 2.2 Neuropsychological testing

Participants were administered four neuropsychological memory tasks: NEUROPSI, 2^nd^ edition (NEUROPSI, 2^nd^ ed.) Logical Memory, the Free and Cued Selective Reminding Test (FCSRT), NEUROPSI, 2^nd^ ed. Word List, and NEUROPSI, 2^nd^ ed. Semi-Complex Figure. The NEUROPSI battery was selected because it was developed especially for use in Latin American individuals, with close regard to the social and cultural characteristics of this population^19^. Specific scores used in analyses included: NEUROPSI Logical Memory Delayed Recall, FCSRT Total Recall, NEUROPSI Word List Delay Free Recall, and NEUROPSI Semi-Complex Figure Delay Total. Raw scores for each measure were converted into z-scores with a mean of 0 and standard deviation of 1 using regression coefficients derived from cognitively unimpaired individuals in the BLAST study. The memory IIV index was created by computing the intraindividual SD across the four memory z-scores, consistent with previous studies^4,6,20,21^. Mean memory performance was calculated as the average of z-scores across the same measures. Participants were included if they had at least two of the four neuropsychological test scores available, although 66 of the 67 participants had all four scores available.

### 2.3 MRI acquisition

Participants completed an MRI scan at Massachusetts General Hospital in Boston. Structural MRI acquisition of the whole head (T1-weighted MPRAGE) was performed on a 3 Tesla Tim Trio system.

The MRI protocol also included diffusion-weighted images using a single shot spin echo planar imaging sequence [repetition time (TR) = 8040 msec, echo time (TE) = 84 msec, flip angle (FA) = 80°, field of view (FOV) = 256 × 256 mm, voxel size = 2.0 × 2.0 × 2.0 mm3, 30 diffusion-sensitizing gradients with a b-value of 700 s/mm2] and 5 non-diffusion -weighted images (b = 0 s/mm2)^22^.

### 2.4 MRI preprocessing and analysis

We used SPM8 for MRI preprocessing and voxel-based morphometry analysis of the anatomical T1-weighted MR images^23^. Briefly, after spatial normalization of all images from native to normalized MNI/ICBM space, images were segmented into GM, white matter, and CSF. Then, the modulated GM images were selected for additional analysis and smoothed using an 8 mm full-width at half-maximum Gaussian kernel. For computational efficiency, particularly for the graph theoretic analyses, we down sampled all neuroimaging data to 6 mm isotropic voxels. Then, we vectorized all individual imaging data and concatenated all subjects in a single voxel-by-subjects matrix for each imaging modality^24^. Bilateral hippocampal volume was normalized to the estimated total intracranial volume.

Standard pre-processing for diffusion-weighted data was performed using FSL (v5.0.9; University of Oxford, UK) and AFNI (v17.2.10)^25^. Data were corrected for head movement and eddy currents, skull-stripped, masked, and fit for diffusion tensors. Voxel-wise scalar maps were thresholded to exclude FA values below 0.2, normalized to the MNI152 template, and skeletonized for group-level Tract-Based Spatial Statistics (TBSS)^26^. Hippocampal white matter integrity was assessed using mean FA extracted from the left and right cingulum-hippocampus using the JHU ICBM white matter atlas (25% probability threshold)^27^. Atlas-defined tract masks were intersected with the TBSS skeleton mask to restrict extraction to skeleton voxels, and mean FA values were calculated within each mask using FSL’s fslmeants^28^.

### 2.5 Statistical analysis

Multiple linear regression models, adjusting for age, sex, education, cognitive status, and mean memory performance, were run to assess associations between memory IIV and hippocampal integrity, measured by bilateral hippocampal volume and fractional anisotropy in the hippocampus-cingulum tract. In subsequent analyses, mean memory performance replaced memory IIV as the main outcome of interest and multiple linear regressions were adjusted for age, sex, education, and cognitive status.

## 3. Results

### 3.1 Sample characteristics

The sample included 67 participants with complete MRI and neuropsychological data available. Participants had a mean age of 65 years (SD = 6.94), 11.49 years of education (SD = 4.93), and 81% (n = 54) of the sample was female. Seven individuals were classified as cognitively impaired based on total MMSE scores. Demographic characteristics are presented in Table 1.

**Table 1.** Demographic data of sample.

| **<u>Demographics</u>** | Mean (Standard Deviation) |
| --- | --- |
| Age (years) | 65.00 (6.94) |
| Education (years) | 11.49 (4.93) |
| Female, n (%) | 54 (81%) |
| Cognitively Unimpaired, n (%) | 60 (90%) |
| Mini Mental State Examination | 26.93 (2.55) |
| Free and Cued Selective Reminding Test Total Recall | 46.11 (5.14) |
| NEUROPSI Logical Memory Delayed Recall | 6.13 (3.03) |
| NEUROPSI Word List Delay Free Recall | 5.85 (2.79) |
| NEUROPSI Semi-Complex Figure Delay Total | 7.20 (2.55) |

### 3.2 Memory IIV, memory performance and demographic characteristics

Memory IIV was not significantly associated with age, sex, education, cognitive status, or mean memory performance. Lower mean memory performance was associated with older age (*β*=-0.27, Adj. R^2^= 0.34, *p*=.02) but not sex, education, or cognitive status. There was no multicollinearity among predictors (all VIFs <2). Results are presented in Tables 2 and 3.

**Table 2.** Associations of memory intraindividual variability (IIV)

| **<u>Variable</u>** | $\beta$ | SE | t | p-value |
| --- | --- | --- | --- | --- |
| Age (years) | -0.222 | 0.006 | -1.583 | 0.119 |
| Education (years) | -0.124 | 0.008 | -0.870 | 0.388 |
| Sex (Male) | -0.016 | 0.093 | -0.128 | 0.899 |
| Cognitive Status (Cognitively Unimpaired) | 0.018 | 0.144 | 0.123 | 0.903 |
| Mean Memory Performance | -0.094 | 0.052 | -1.807 | 0.076 |
| Bilateral Hippocampal Volume | 0.028 | 76.749 | 0.201 | 0.841 |
| FA Hippocampus-Cingulum Tract | -0.335 | 0.551 | -2.748 | **0.008** |

**Table 3.** Associations of mean memory performance.

| **<u>Variable</u>** | **$\beta$** | **SE** | **t** | **p-value** |
| --- | --- | --- | --- | --- |
| Age (years) | -0.266 | 0.014 | -2.420 | **0.018** |
| Education (years) | 0.203 | 0.020 | 1.785 | 0.079 |
| Sex (Male) | -0.053 | 0.225 | -0.520 | 0.605 |
| Cognitive Status (Cognitively Unimpaired) | 0.326 | 0.330 | 2.798 | **0.007** |
| Bilateral Hippocampal Volume | 0.236 | 172.522 | 2.234 | **0.029** |
| FA Hippocampus-Cingulum Tract | -0.068 | 1.411 | -0.645 | 0.522 |

### 3.3 Associations between memory performance and neuroimaging outcomes

Memory IIV was not related to bilateral hippocampal volume (*β*=0.03, Adj. R^2^= 0.004, *p*=0.84). Larger hippocampal volume was associated with higher mean memory performance (*β*=0.24, Adj. R^2^= 0.38, *p*=.03). Memory IIV was associated with lower fractional anisotropy in the hippocampus-cingulum tract (*β*=-.34, Adj. R^2^= 0.11, *p*<.01), while mean memory performance was not (*β*=-0.07, Adj. R^2^= 0.34, *p*=.52). There was no multicollinearity among predictors (all VIFs <2). Results are presented in Tables 2 and 3 and Figure 1.

**Figure 1.**
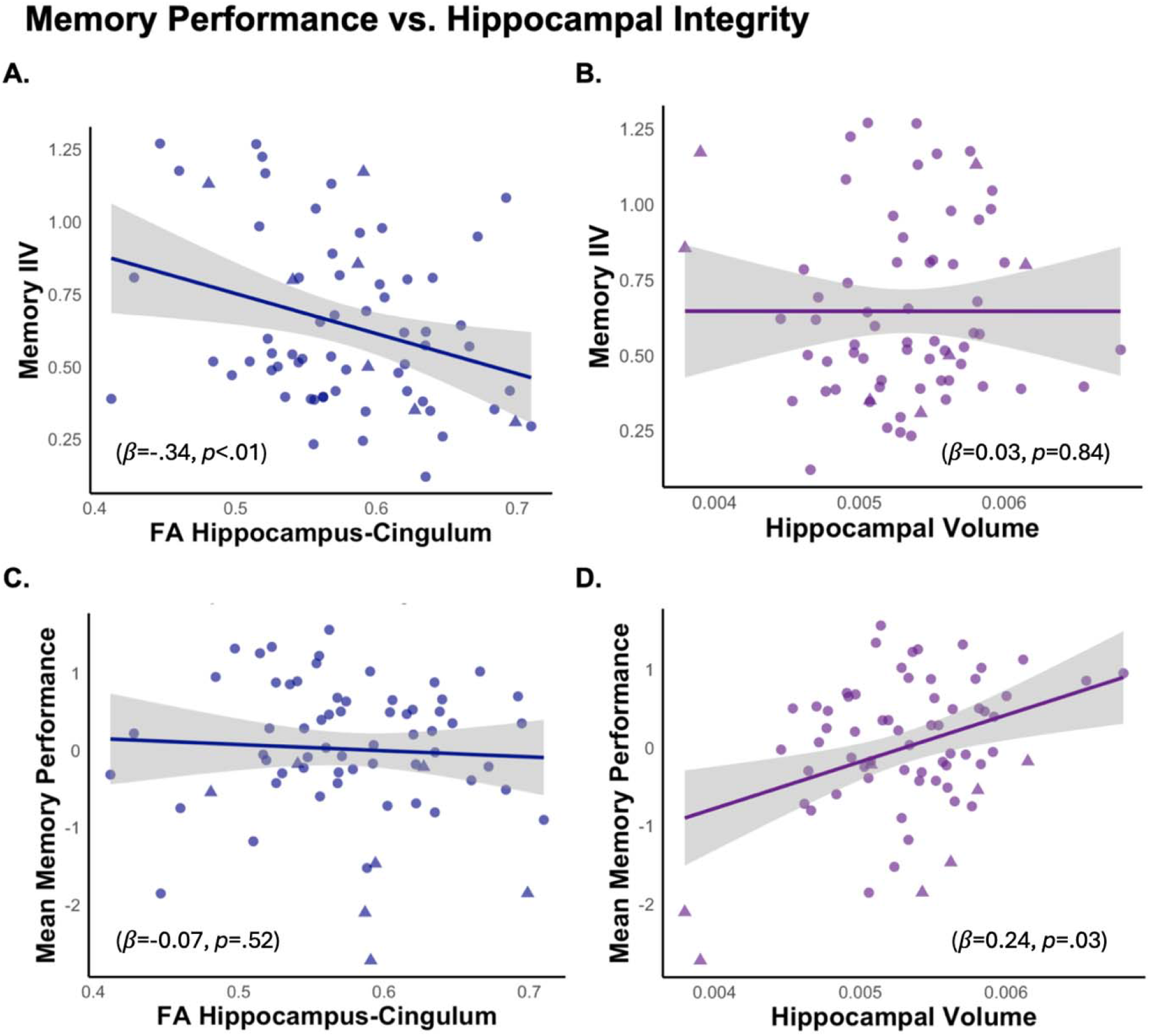
Memory Performance and Hippocampal Integrity. Figure caption: Linear regression results illustrate associations among memory performance (i.e., memory IIV and mean memory performance) and neuroimaging measures (i.e., FA hippocampus-cingulum and hippocampal integrity). Cognitively unimpaired individuals are represented by circles, and cognitively impaired individuals are represented by triangles. IIV = intraindividual variability; FA = fractional anisotropy.

## 4. Discussion

In this study, we examined associations among memory, demographic characteristics, and markers of hippocampal integrity in a cohort of older Latino adults. Memory IIV was unrelated to age, sex, education, cognitive status, and mean memory performance, while lower mean memory performance was associated with older age and cognitive impairment. Importantly, memory IIV and mean memory performance showed distinct neuroanatomical correlates. Greater memory IIV was associated with lower fractional anisotropy in the hippocampus-cingulum tract but not hippocampal volume, while mean memory performance was associated with hippocampal volume but not white matter integrity. Together, these findings suggest that memory IIV and mean memory performance may capture different aspects of hippocampal dysfunction and potentially reflect distinct stages of neurodegenerative change.

The differential associations observed for memory IIV and mean memory performance may provide insight into the neural mechanisms underlying these cognitive processes. Memory IIV appeared to be more sensitive to disruptions in white matter connectivity, suggesting that greater variability across memory tasks may be a result of reduced efficiency of communication among brain regions involved in memory processing, even when core hippocampal functions remain relatively preserved. In contrast, mean memory performance may more directly reflect the integrity of hippocampal gray matter structure that support memory encoding, consolidation, and retrieval. The findings also lend support to the potential utility of memory IIV as a marker of early white matter disruption in individuals at increased risk for dementia.

Our findings both align with and extend previous research on memory IIV. Previous studies have found associations between memory IIV and cognitive status^15^; however, we did not observe that relationship. Differences in study populations may explain discrepancy in findings, and our relatively small number of cognitively impaired individual may have limited our ability to detect such effects. Consistent with prior work demonstrating that cognitive IIV is sensitive to early white matter changes^4^, greater memory IIV in our sample was associated with reduced white matter integrity. In contrast, although some studies have reported associations between cognitive variability and hippocampal volume^6^, memory IIV was unrelated to hippocampal volume in this present sample. Together, these findings suggest that within-domain memory IIV may provide information that is distinct from traditional tasks in Latino populations. Given its noninvasive and scalable nature, memory IIV may represent a promising approach for identifying early white matter neurodegeneration, particularly of the cingulum bundle. However, more work is needed to solidify the clinical utility of both within-domain and across domain IIV for underrepresented populations.

Several strengths and limitations should be considered when interpretating these findings. A major strength of the study is the focus on Spanish-speaking Latino adults, a population that remains underrepresented in dementia research despite being at increased risk for cognitive decline and dementia. We also have access to several Spanish-language tests of memory including encoding and consolidation tasks. At the same time, the cross-sectional design limits our ability to establish causal relations or determine relevant temporal cascades. The modest sample size may be limited in its power to detect smaller effect sizes for relationships between cognition and neuroimaging measures. In addition, our investigation focused on memory IIV, and it remains unclear whether similar relationships would be observed within or across other cognitive domains. Another limitation is the absence of vascular risk factors as covariates in our analyses, as vascular health is known to contribute to brain structural integrity, especially as it relates to white matter. Future longitudinal studies are needed to determine whether higher baseline memory IIV predicts subsequent cognitive decline, accelerated neurodegeneration or conversion to dementia in Latino older adults. Further research should also examine whether cognitive IIV is sensitive to early ADRD biomarkers, including fluid-based markers of pathology. Such work will be important for clarifying the clinical utility of both within domain and across-domain IIV measures and for evaluating their potential role in the early detection of neurodegenerative diseases in this population.

Overall, the present findings suggest that memory IIV may be more sensitive to hippocampal white matter integrity, while mean memory performance appears more closely linked to hippocampal volume. These differential associations suggest that memory IIV and mean performance may capture distinct aspects of neurodegenerative processes and may reflect different stages of disease progression. Importantly, our results highlight the value of considering variability in cognitive performance alongside traditional mean scores, as IIV may provide unique information about underlying neural integrity. This work adds to growing literature supporting IIV as a tool and potentially sensitive marker for early detection of neurodegeneration and dementia risk.

## Consent Statement

All participants provided written informed consent before participating and were compensated. The study was approved by the Massachusetts General Hospital (Boston, U.S.A.) local institutional review board.

## Data availability statement

Anonymized clinical and imaging data are available upon request, subject to an internal review by Y.T.Q. to ensure that the participants’ anonymity and confidentiality are protected. Data requests will be considered based on a proposal review, and completion of a data sharing agreement, in accordance with Mass General Brigham institutional guidelines. Please submit data requests to Y.T.Q.

## Acknowledgments

The authors express their gratitude to the BLAST research participants for generously contributing their time and effort.

## CRediT authorship contribution statement

Amanda I. Gonzalez: Writing – review & editing; Writing – original draft; Methodology; Formal analysis; Investigation; Conceptualization; Visualization. Catarina Tristão-Pereira: Writing – review & editing; Investigation; Methodology. Lusiana Martinez: Writing – review & editing; Investigation; Data curation. Nadeshka J. Ramirez: Writing – review & editing; Investigation; Data curation. George Aquilino Silva: Writing – review & editing; Investigation; Data curation. Jorge Alcina: Writing – review & editing; Investigation. Jairo E. Martinez: Writing – review & editing; Investigation. Averi Giudicessi: Writing – review & editing; Investigation. Vivian Ku: Writing – review & editing; Investigation. Meredith C. Rowe: Writing – review & editing; Investigation. Liliana A. Ramirez-Gomez: Writing – review & editing; Investigation. Daniel G. Saldana – review & editing; Investigation. Yakeel T. Quiroz: Writing – review & editing; Conceptualization; Investigation; Supervision; Funding acquisition.

